# The association between household use of unclean cooking fuels and depression symptoms among older adults in India: a cross-sectional study

**DOI:** 10.64898/2026.04.13.26350749

**Authors:** K. Mohsini, GR. Gore-Langton, SD. Rathod, KE. Mansfield, C. Warren-Gash

## Abstract

**Aims:** Indoor air pollution resulting from combustion of unclean cooking fuels has been linked to adverse health outcomes, but evidence regarding its association with mental health in low-and middle-income countries remains limited. We investigated the association between household use of unclean cooking fuels, as a proxy for indoor air pollution, and depression symptoms among adults aged 45 years and older in India, and assessed effect modification by age, sex, caste, and rural/urban residence.

**Methods:** We conducted a cross-sectional analysis of the first wave (2017–2018) of data from the Longitudinal Aging Study in India (LASI), a nationally representative survey of adults aged ≥45 years. Cooking fuel type was classified as clean or unclean, and depression symptoms were assessed using the 10-item Centre for Epidemiologic Studies Depression (CES-D-10) scale. We used logistic regression to estimate odds ratios for depression symptoms, and linear regression to compare mean CES-D-10 scores by cooking fuel type, adjusting for sociodemographic and housing characteristics.

**Results:** We included 62,650 respondents. Median age was 57 years (IQR: 50–65), 46.7% were women, 47.6% reported using unclean cooking fuels, and 27.6% screened positive on the CESD-10. After adjusting for sociodemographic and housing characteristics, use of unclean cooking fuels was associated with higher odds of screening positive on the CESD-10 (aOR: 1.08; 95% CI: 1.02, 1.15), and higher mean CES-D-10 scores (adjusted mean difference: 0.34; 95% CI: 0.24, 0.44). The association was more pronounced among individuals living in urban areas (aOR: 1.36; 95% CI: 1.21, 1.53).

**Conclusion:** Use of unclean cooking fuels was associated with depression symptoms among older adults in India, and especially among those living in urban areas.

## Introduction

In India, rapid demographic changes mean that mortality rates are falling, older people are living longer, and population ageing is occurring at an accelerated rate, concurrent with increased prevalence and early onset of multiple chronic diseases [1, 2]. Projections suggest that those aged 60 and above will constitute 19.5% of India’s population by 2050, accounting for 319 million elderly people [3], presenting a major challenge given low public investment in healthcare and welfare systems. Among elderly people, there is an escalating burden of mental health conditions [4], which are risk factors for poor brain health outcomes such as cognitive impairment and neurodegenerative disease [5]. The prevalence of depressive disorders increases with age in India, reaching 6.5% among those aged 60 and older [6]; prevalence is higher in females than in males, starting at 45 years [7]. Lack of awareness and sensitivity regarding mental health conditions means those experiencing depression symptoms rarely seek diagnosis and treatment, which is compounded by inadequate evidence-based care, and poorly implemented and unequally distributed mental health services [8]. The treatment gap for depression (proportion of people with depression who meet criteria for care but do not receive treatment) is estimated to be 79.1% [9, 10], with only 0.3 psychiatrists per 100,000 people in India [11]. Identifying modifiable risk factors for depression is crucial to preventing and addressing the burden.

Indoor air pollution is generated by incomplete combustion of unclean fuels – such as biomass (wood, crop residues, animal dung, and charcoal), coal, and kerosene – typically used for cooking, often powering poorly ventilated and inefficient stoves, ovens or open fires within the home [12]. An estimated 28.9% of India’s overall population and 56.8% of the rural population rely on unclean cooking fuels [13]. Additionally, an estimated 0.61 million deaths were attributable to household air pollution in 2019, making up 6.5% of total deaths in India [14, 15]. Polluting compounds such as black carbon, PM_2.5_ (fine particulate matter with diameter ≤2.5μm), carbon monoxide, and sulphur dioxide can penetrate the lungs, enter the bloodstream, cross the blood-brain barrier, and negatively affect brain health by causing inflammation [16, 17]. Mechanistic studies have shown that exposure to air pollutants is associated with neuroinflammation, oxidative stress, and cerebrovascular damage [18, 19]. Neuroinflammation induced by exposure to PM_2.5_ is hypothesised to alter the physiological expression of clock genes, dysregulating circadian rhythms and triggering low mood [20–22]. Furthermore, exposure to PM_2.5_ has been associated with activation of the hypothalamus–pituitary–adrenal axis and prolonged secretion of the stress hormone cortisol [23], commonly implicated in depression aetiology [24].

Longitudinal evidence from China suggests that exposure to unclean cooking fuels is associated with an increased risk of depression [25–28]. One systematic review of cohort studies reported a 23% higher risk of depression among users of polluting cooking fuels (RR: 1.23; 95%□CI: 1.16, 1.31) [29]. These findings may not be generalisable to India, where cooking fuel choices differ, mental healthcare infrastructure is weaker and a larger proportion of the population lives below the international poverty line [30]. To our knowledge, two previous studies have examined the hypothesised association between unclean cooking fuel and depression symptoms in India, where it is yet to be investigated longitudinally. A cross-sectional study in rural West Bengal found that cooking with biomass is associated with higher odds of mild-to-severe depression symptoms (OR: 1.67; 95% CI: 1.18, 2.95) in 1756 pre-menopausal women [31], although this was not a population-based sample, limiting its generalisability. Another study restricted to adults aged 60 and above from Wave 1 of the Longitudinal Aging Study in India (LASI), who comprised less than half of the LASI study population, found a modest association between polluting fuel use and depression symptoms (OR: 1.09; 95% CI: 1.03, 1.16) [32]. However, it remains unclear whether this association is observed among adults below age 60 or varies across age groups. To address these gaps, we conducted a cross-sectional analysis using nationally representative data LASI data to investigate the association between household use of unclean cooking fuels and depression symptoms, including the full sample of adults aged 45 and above.

## Materials and methods

### Study design and population

We used data from LASI, a nationally representative study of adults aged 45 and older and their spouses (regardless of age) from 35 of the 36 states and union territories of India (excluding Sikkim) [33]. For LASI Wave 1, the sampling frame was India’s 2011 Census, which only included community-dwelling, household populations. Individuals living in institutional living arrangements were not included.

LASI adopted a multistage stratified area probability cluster sampling design, entailing three-stage sampling design in rural areas and four-stage sampling design in urban areas. In each state, sub-districts (tehsils/talukas) were identified as primary sampling units (PSUs). In urban areas, wards were selected from each PSU. From each ward, one census enumeration block (CEB) was selected as the secondary sampling unit (SSU), from which households with at least one member aged 45 or above were eligible to be selected via systematic random selection. In rural areas, villages were selected as SSUs. Overall, household response rate was 95.8% and individual response rate was 87.3%, with 72,250 adults participating. We removed observations for individuals below age 45 years, those missing data on exposure or outcome, and those for whom a proxy interview was conducted (those incapable of giving a full interview).

### Data collection

Data were collected by trained field investigators conducting face-to-face interviews and direct health measures between April 2017 and December 2018. The LASI instrument, consisting of individual, household, and community survey schedules, was translated into 16 regional languages. Informed consent was obtained from eligible households and individuals in their preferred languages. The consent form was read to respondents who were unable to read, and they were asked to provide a signature or inked fingerprint. Individual surveys collected information on demographics, functional health, mental health (cognition and depression symptoms), work, retirement, and pensions, healthcare services utilisation, family and social networks, and social welfare utilisation. Questions were asked on household traits such as physical environment, food consumption, income, and expenditure. The Harmonised LASI dataset is publicly available in anonymised, de-identified form and was accessed for research purposes between July and September 2024. The authors did not have access to information that could identify individual participants at any stage of the analysis.

### Exposure

Our exposure was self-reported combustion of unclean cooking fuel as the main cooking fuel in the home, which served as a proxy for exposure to indoor air pollution. Based on previous literature the following were classified as unclean cooking fuels (as burning them produces large amounts of smoke, which is a mixture of gases, particulate matter, and organic chemicals that are harmful to health): wood, shrub, crop residues, dung cake, charcoal, coal, lignite, and kerosene. Liquid petroleum gas (LPG), biogas, and electricity were classified as clean cooking fuels.

### Outcome

Our outcome, depression symptoms (as a proxy for depression), was measured using the 10-item Centre for Epidemiologic Studies Depression (CES-D-10) scale, which has been validated in multiple settings, including among older adults in India [34–36]. The CES-D-10 includes seven negative symptoms (trouble concentrating, feeling depressed, low energy, fear of something, feeling alone, bothered by things, and everything is an effort), and three positive symptoms (feeling happy, hopeful, and satisfied). The four response options reflect the frequency of symptoms experienced in the past week prior to interview: “rarely or never” (<1 day), “sometimes” (1 or 2 days), “often” (3 or 4 days), and “most or all of the time” (5 to 7 days). An intermediary variable assigns dichotomous scores; for negative symptoms, “rarely or never” and “sometimes” are scored 0, and “often” and “most or all of the time” categories are scored 1 (S1 Table). Scoring is reversed for positive symptoms. Overall scores ranged from 0 to 10. The binary outcome “presence of depression symptoms” was defined as a score of 4 or above, which has a sensitivity of 97.7% and a specificity of 79.1% for screening depression symptoms in community-dwelling older adults in India, according to the validation study [34]. For our sample, the CES-D-10 scale demonstrated good internal consistency (Cronbach’s α = 0.80), aligning with the validation study.

### Covariates

We considered the following variables as potential confounders: categorical age, sex, marital status, rural/urban location, covariates under the construct of socioeconomic vulnerability (caste, literacy, education, employment, and whether the household lives above or below the international poverty line), and covariates under the construct of housing quality (whether the household has improved sanitation, permanent/temporary housing materials, separate bedrooms, electricity, or a water facility inside own dwelling). Details of the conceptual framework developed to guide covariate selection are provided in S1 Fig and S2 Table (which includes a description of and rationale behind each covariate included in our model and its categorisation). We considered the caste categories Scheduled Castes (SC), Scheduled Tribes (ST), Other Backward Castes (OBC), and no/other caste, comprising other groups with or without caste affiliation. SC and ST are the most historically disadvantaged castes, who experience exclusion from social and economic opportunities, as well as disproportionate exposure to pollutants, toxins, and morbidity [37, 38]. OBC face marginalisation but are slightly better off, while those of no/other caste are the most privileged.

### Statistical analysis

We survey-weighted data at the individual level using the design weight provided in the Harmonised LASI data to account for unequal selection probabilities of households, and therefore individuals, differential non-response rates, and to bring the sample in line with key sociodemographic variables in the reference population. We used Stata 18 to conduct all analyses [39]. We have included our Stata analysis code in the Supporting Information.

We summarised non-survey-weighted and survey-weighted descriptive characteristics of individuals using clean or unclean cooking fuels, presenting frequency (n [%]) for categorical characteristics and median (interquartile range [IQR]) for continuous variables. We also summarised differences in characteristics between individuals included and excluded (due to proxy interview, missing data on the exposure or outcome, or being below age 45 years) in the analyses. All subsequent analyses were adjusted for the complex survey design.

We analysed the outcome as a binary variable (presence/absence of depression symptoms) using multivariate logistic regression to estimate odds ratios (OR) and 95% confidence intervals (95% CI) to assess if those using unclean cooking fuels had higher odds of screening positive for depression symptoms compared to those using clean cooking fuels. Our adjusted model controlled for confounding variables collectively representing the constructs of socioeconomic vulnerability (caste, literacy, education, employment, and above/below international poverty line) and housing quality (improved sanitation, housing materials, electricity, water facility inside own dwelling, and separate rooms), and rural/urban residence.

We also analysed the outcome as a continuous measure, using multivariate linear regression to estimate the mean difference of CES-D-10 score between users of clean and unclean cooking fuels, controlling for the same confounders (as the logistic regression analysis). We assessed linear modelling assumptions using residual diagnostics.

### Secondary analyses

We investigated whether the association between unclean cooking fuel and presence/absence of depression symptoms as a binary outcome was modified by: (1) age group (45-59 years, 60-74 years, 75+ years); (2) sex (male/female); (3) urban/rural residence; and (4) caste (SC, ST, OBC, no/other). We included these potential effect modifiers as interaction terms in multivariable logistic regression models, adjusted for all other confounders. Evidence of effect modification was assessed using post-estimation contrast tests that account for the complex survey design, with p<0.005 considered as a meaningful indicator of heterogeneity.

## Results

### Statistical analysis

We identified a total of 72,262 individuals who participated in interviews, of whom 65,575 were aged 45 years and over. We excluded 694 participants who had proxy interviews, as they were incapable (cognitively or physically) of giving a full interview, and those missing data on exposure or outcome, resulting in an analysis sample of 62,650 respondents (S2 Fig). The baseline characteristics of those included in final, fully adjusted regression models were broadly similar to those excluded (S3 Table).

Accounting for survey weighting, median age was 57 years (IQR: 50–65), 46.7% of respondents were women, 77.5% were married, and 68.2% lived in rural areas. In total, 18.4% of respondents lived below the poverty line, 73.1% belonged to SC/ST/OBC castes, 53.0% were illiterate, and 62.3% had received no education. Regarding household characteristics, 91.9% had electricity, 64.9% had improved sanitation, 66.1% owned a water facility inside their dwelling, 45.1% were made of temporary housing materials, and 80.1% had separate bedrooms. Forty-seven percent (47.6%) of respondents reported using unclean cooking fuels. Use of LPG, a cleaner alternative, was substantially higher in urban households (88.3%) than in rural households (34.9%). In rural areas, the most common primary cooking fuel was wood or shrubs (50.8%), followed by dung cake (6.9%) and charcoal, lignite or coal (3.5%) [40]. The proportion with depression symptoms was 27.6%. Median CES-D-10 score was 9 [IQR: 7–12].

Respondents using unclean cooking fuels were more likely than those using clean fuels to be illiterate (68.3% compared to 39.1%), have no formal education (77.8% compared to 48.1%), live below the international poverty line (28.2% compared to 9.5%), and live in rural areas (91.4% compared to 47.0%) (Table 1). Unclean cooking fuel users were less likely than clean fuel users to have improved sanitation, permanent housing materials, separate bedrooms, electricity, or own a water facility inside their dwelling. The prevalence of depression symptoms was higher among unclean cooking fuel users at 30.6% compared to clean cooking fuel users at 24.9%.

**Table 1:** Characteristics of LASI respondents by use of clean or unclean cooking fuel (N=63,597), India, 2017-2018.

|  | Uses clean cooking fuel (N=34,230) |  | Uses unclean cooking fuel (N=29,367) |  |
| --- | --- | --- | --- | --- |
|  | Unweighted frequency | Weighted % | Unweighted frequency | Weighted % |
| **Individual characteristics** |  |  |  |  |
| **Age (years) (median [IQR])** | 58.0 [50.0–66.0] | 57.0 [50.0–65.0] | 59.0 [51.0–67.0] | 58.0 [51.0–65.0] |
| **Age (years)** |  |  |  |  |
| 45–59 | 18,375 | 58.4 | 14,989 | 56.0 |
| 60–74 | 12,605 | 33.4 | 11,349 | 35.1 |
| 75+ | 3,249 | 8.2 | 3,027 | 8.9 |
| Missing | <50 | - | <50 | - |
| **Sex** |  |  |  |  |
| Male | 15,953 | 53.6 | 13,634 | 53.3 |
| Female | 18,277 | 46.4 | 15,733 | 46.7 |
| **Caste** |  |  |  |  |
| Scheduled Castes | 5,010 | 16.1 | 5,643 | 23.9 |
| Scheduled Tribes | 3,374 | 4.0 | 7,694 | 13.8 |
| Other Backward Castes | 13,543 | 46.5 | 10,411 | 42.6 |
| No caste/other | 11,978 | 33.4 | 5,464 | 19.7 |
| Missing | 325 | - | 155 | - |
| **Literacy** |  |  |  |  |
| Literate | >22,078 | 61.0 | >10,751 | 31.7 |
| Illiterate | 12,102 | 39.0 | 18,566 | 68.3 |
| Missing | <50 (<0.1) | - | <50 | - |
| **Education** |  |  |  |  |
| None/below primary | >15,3121 | 48.1 | 21,713 | 77.8 |
| Primary | 5,014 | 13.6 | 3,359 | 9.4 |
| Middle school | 3,964 | 8.9 | 2,138 | 5.8 |
| Secondary | 6,594 | 19.5 | 1,833 | 5.9 |
| Diploma/grad/post-grad | 3,296 | 9.9 | 324 | 1.1 |
| *Missing* | <50 | - | 0 | - |
| **Employment** |  |  |  |  |
| Yes | >15,381 | 52.5 | >16,185 | 60.1 |
| No | 18,799 | 47.5 | 13,132 | 39.9 |
| *Missing* | <50 | - | <50 | - |
| **Marital status** |  |  |  |  |
| Married/partnered | 25,991 | 77.8 | 21,998 | 77.2 |
| Separated/divorced/widow | 8,237 | 22.2 | 7,369 | 22.8 |
| *Missing* | <50 | - | 0 | - |
| **Weekly contact with family/friends** |  |  |  |  |
| Yes | 4,707 | 13.4 | 3,858 | 10.2 |
| No | 28,996 | 86.6 | 25,194 | 89.8 |
| *Missing* | 527 | - | 315 | - |
| **BMI\*** |  |  |  |  |
| Underweight (BMI<18.5) | 3,213 | 11.5 | 7,587 | 30.4 |
| Normal (18.5≤BMI<23) | 9,771 | 33.0 | 11,887 | 43.5 |
| Overweight (23≤BMI<25) | 5,314 | 17.1 | 3,451 | 11.9 |
| Obese (BMI≥25) | 12,577 | 38.4 | 4,304 | 14.2 |
| *Missing* | 3,355 | - | 2,138 | - |
| **Alcohol consumption** |  |  |  |  |
| Never | 30,953 | 90.9 | 25,665 | 88.2 |
| Infrequent | 1,728 | 5.4 | 2,080 | 7.0 |
| Frequent | 1,245 | 3.7 | 1,455 | 4.8 |
| *Missing* | 304 | - | 167 | - |
| **Smoker status** |  |  |  |  |
| Never | 28,815 | 84.0 | 22,729 | 75.7 |
| Ever | 5,100 | 16.0 | 6,467 | 24.3 |
| *Missing* | 315 | - | 171 | - |
| **Medical visits in previous year** |  |  |  |  |
| Yes | 18,703 | 57.2 | 14,912 | 57.1 |
| No | 15,123 | 42.8 | 14,235 | 42.9 |
| *Missing* | 404 | - | 220 | - |
| **Household characteristics** |  |  |  |  |
| **International poverty line\*\*** |  |  |  |  |
| Above | 31,451 | 90.5 | >21,330 | 71.8 |
| At/below | 2,779 | 9.5 | 7,987 | 28.2 |
| *Missing* | 0 | - | <50 | - |
| **Rural/urban residence** |  |  |  |  |
| Rural | 15,152 | 47.0 | 26,165 | 91.4 |
| Urban | 19,078 | 53.0 | 3,202 | 8.6 |
| **Improved sanitation\*\*\*** |  |  |  |  |
| Yes | >28,450 | 80.1 | >17,372 | 48.0 |
| No | 5,712 | 19.9 | 11,945 | 52.0 |
| *Missing* | <50 | - | <50 | - |
| **Water facility inside own dwelling** |  |  |  |  |
| Yes | >26,892 | 75.0 | >17,370 | 56.4 |
| No | 7,288 | 25.0 | 11,947 | 43.6 |
| *Missing* | <50 | - | <50 | - |
| **Housing materials (pucca/kutchra)** |  |  |  |  |
| Permanent (pucca) | 24,781 | 73.4 | >9,681 | 34.6 |
| Temporary (kutchra) | 9,373 | 26.6 | 19,636 | 65.4 |
| *Missing* | 76 | - | <50 | - |
| **Separate bedrooms** |  |  |  |  |
| Yes | >29,713 | 84.6 | >23,180 | 75.1 |
| No | 4,467 | 15.4 | 6,137 | 24.9 |
| *Missing* | <50 | - | <50 | - |
| **Electricity** |  |  |  |  |
| Yes | 33,857 | 98.4 | 25,941 | 84.7 |
| No | 373 | 1.6 | 3,426 | 15.3 |

\*We recoded BMI into four categories of underweight (BMI<18.5), normal (18.5≤BMI<23), overweight (23≤BMI<25), and obese (BMI≥25), based upon classification criteria specific to South Asian ethnic groups, recommended by several guidelines for prevention and management of obesity-related NCDs [41]
\*\*International poverty line is defined by the World Bank and currently set at \$1.90 per person per day in 2011 purchasing power parity (PPP) dollars
\*\*\*Improved sanitation includes flush or pour flush toilet connected to piped sewage system, septic tank or pit latrine, or twin pit/composting toilet or pit latrine, and indicates that household does not share their toilet facility with any other households

After correction for survey-weighting, having fully adjusted for all available potential confounders, those who reported using unclean cooking fuels were 8% more likely to experience depression symptoms (OR: 1.08; 95% CI: 1.02,1.15; p = 0.007) compared to those using clean cooking fuels.

In linear regression analyses, the residuals versus fitted values plot exhibited random scatter with no discernible systematic patterns, supporting linearity and homoscedasticity. The Q-Q plot suggested minor deviations from normality around the tail, which we concluded were unlikely to bias estimates given the large sample size. In the adjusted linear model, unclean cooking fuel users had a higher mean depression symptom score by 0.34 points (95% CI: 0.24, 0.44; p<0.001) (Table 2).

**Table 2:** Association between unclean cooking fuel use and depression symptoms among older adults in India, 2017-2018 (N=62,650).

| Probable depression |  |  |
| --- | --- | --- |
|  | Weighted number of depression events in individuals using clean/unclean cooking fuel | OR (95% CI**) |
| Clean | n=8536/33,653 | 1 (ref) |
| Unclean* | n=9540/28,997 | 1.08 (1.02, 1.15) |
| Mean difference in depression symptom score |  |  |
|  | Weighted number of depression events in individuals using clean/unclean cooking fuel | Beta (95% CI**) |
| Clean | n=8536/33,653 | 0 (ref) |
| Unclean* | n=9540/28,997 | 0.34 (0.24, 0.44) |

\*Adjusted for adjusted for age, sex, marital status, caste, education, literacy, employment, poverty, rural/urban residence, improved sanitation, housing materials, electricity, water facility inside own dwelling, and separate rooms
\*\*CI = confidence interval – obtained using robust standard errors

We did not find evidence of effect modification by categorical age or sex (Table 3).

**Table 3:**
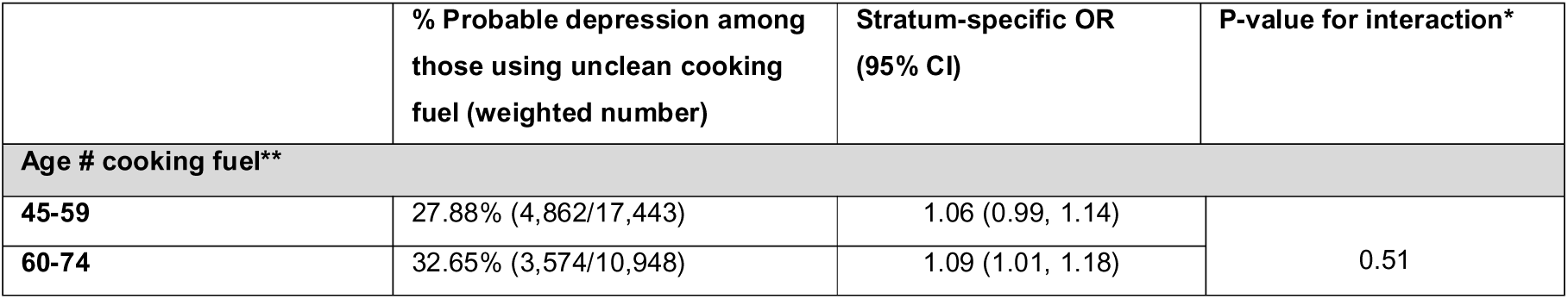
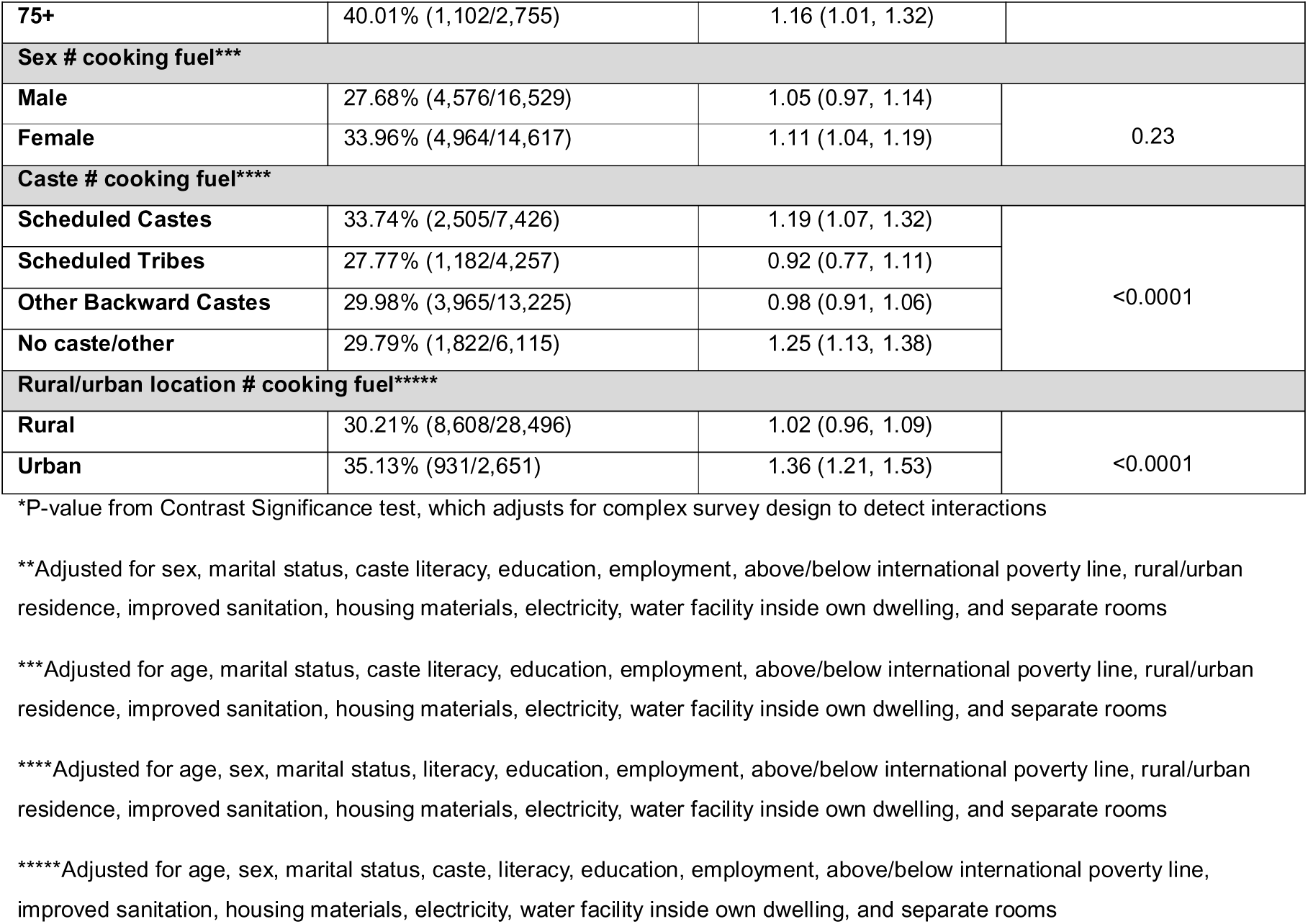
Interaction effects of age, sex, caste, and rural/urban location on the association between unclean cooking fuel use and odds of probable depression among adults aged 45 and older.

|  | % Probable depression among those using unclean cooking fuel (weighted number) | Stratum-specific OR (95% CI) | P-value for interaction* |
| --- | --- | --- | --- |
| Age # cooking fuel** |  |  |  |
| 45-59 | 27.88% (4,862/17,443) | 1.06 (0.99, 1.14) | 0.51 |
| 60-74 | 32.65% (3,574/10,948) | 1.09 (1.01, 1.18) |  |
| 75+ | 40.01% (1,102/2,755) | 1.16 (1.01, 1.32) |  |
| Sex # cooking fuel*** |  |  |  |
| Male | 27.68% (4,576/16,529) | 1.05 (0.97, 1.14) | 0.23 |
| Female | 33.96% (4,964/14,617) | 1.11 (1.04, 1.19) |  |
| Caste # cooking fuel**** |  |  |  |
| Scheduled Castes | 33.74% (2,505/7,426) | 1.19 (1.07, 1.32) | <0.0001 |
| Scheduled Tribes | 27.77% (1,182/4,257) | 0.92 (0.77, 1.11) |  |
| Other Backward Castes | 29.98% (3,965/13,225) | 0.98 (0.91, 1.06) |  |
| No caste/other | 29.79% (1,822/6,115) | 1.25 (1.13, 1.38) |  |
| Rural/urban location # cooking fuel***** |  |  |  |
| Rural | 30.21% (8,608/28,496) | 1.02 (0.96, 1.09) | <0.0001 |
| Urban | 35.13% (931/2,651) | 1.36 (1.21, 1.53) |  |

\*P-value from Contrast Significance test, which adjusts for complex survey design to detect interactions
\*\*Adjusted for sex, marital status, caste literacy, education, employment, above/below international poverty line, rural/urban residence, improved sanitation, housing materials, electricity, water facility inside own dwelling, and separate rooms
\*\*\*Adjusted for age, marital status, caste literacy, education, employment, above/below international poverty line, rural/urban residence, improved sanitation, housing materials, electricity, water facility inside own dwelling, and separate rooms
\*\*\*\*Adjusted for age, sex, marital status, literacy, education, employment, above/below international poverty line, rural/urban residence, improved sanitation, housing materials, electricity, water facility inside own dwelling, and separate rooms
\*\*\*\*\*Adjusted for age, sex, marital status, caste, literacy, education, employment, above/below international poverty line, improved sanitation, housing materials, electricity, water facility inside own dwelling, and separate rooms

However, individuals living in urban areas had higher odds of depression symptoms associated with unclean cooking fuel (OR: 1.36; 95% CI: 1.21, 1.53), whereas no evidence of an association was observed in rural areas (OR: 1.02; 95% CI: 0.96, 1.09). Evidence of effect modification by caste was observed (p-value for interaction <0.0001 from a contrast significance test accounting for complex survey design). The association between unclean cooking fuel use and depression symptoms was strongest among respondents of no/other caste (OR: 1.25; 95% CI: 1.13, 1.38) and among Scheduled Castes (OR: 1.19; 95% CI: 1.07, 1.32). In contrast, no clear association was observed among Scheduled Tribes (OR: 0.92; 95% CI: 0.77, 1.11) or Other Backward Castes (OR: 0.98; 95% CI: 0.91, 1.06).

## Discussion and conclusion

In this nationally representative cross-sectional study of adults aged 45 and above in India, we found evidence that use of unclean cooking fuels was associated with 8% higher odds of depression symptoms. Although the absolute difference in mean depression score was modest (0.34 points), the consistent direction of association indicates that household air pollution may influence mental health outcomes in middle-aged and older adults. The higher prevalence of depression symptoms among unclean fuel users (30.6% compared to 24.9% among clean fuel users) underscores the potential contribution of this modifiable environmental factor to India’s mental health burden. The association appeared strongest among urban residents, possibly reflecting residual confounding by unmeasured environmental or housing characteristics in urban areas.

This study benefits from the large sample size and national representativeness of LASI Wave 1, allowing insights into a population where evidence around the distribution or determinants of depression is still emerging. The use of a sampling frame to derive a large, population-based sample minimised selection bias compared to previous studies in India [31].

Several limitations should be considered when interpreting these findings. Exposure to indoor air pollution was assessed indirectly using self-reported primary cooking fuel type rather than quantitative, directly measured pollutant concentrations. This proxy does not capture intensity, duration, or cumulative exposure to indoor air pollutants, nor does it account for the variability of the exposure-response according to fuel type, stove type, ventilation, kitchen characteristics, or time spent cooking, all factors known to influence household-level PM_2.5_ concentrations (regarded as the best measure of indoor air pollution) [42–44]. For example, use of crop residues as cooking fuel was found to result in exposure to the highest PM_2.5_ concentrations compared to cleaner alternatives, followed by dung, wood, and coal [45]. Additionally, not all household members in homes using unclean cooking fuels are equally exposed, particularly those who do not cook or spend time near the stove, where combustion produces concentrated polluting compounds. Self-reported cooking fuel type could also be subject to recall or social desirability bias, with evidence suggesting overreporting of cleaner technologies in survey settings [46].

Depression symptoms were assessed using the CES-D-10, a validated screening tool rather than a clinical diagnosis. Self-reporting of depression symptoms may be influenced by social stigma regarding mental illness, therefore subject to reporting bias, leading to potential underestimation. Although the LASI survey was translated into 16 regional languages according to standardised WHO translation protocol, the psychometric properties of the CES-D-10 scale were not evaluated in each language/dialect, potentially affecting measurement validity for sensitive mental health questions. Reverse causation is unlikely in this study, as depression symptoms experienced in the preceding week would not plausibly influence choice or reporting of household cooking fuel.

Whilst we adjusted for variables capturing socioeconomic and material conditions, measurement of social class and associating various socioeconomic measures with depression is difficult [47]. This difficulty in measuring socioeconomic measures in the context of depression is rendered more complex in India, where social class and caste intersect in complex ways to determine education level, employment, and income. Accordingly, residual confounding by unmeasured socioeconomic variables such as income and occupational exposure is likely. Furthermore, although we conducted our analyses in accordance with Harmonized LASI guidance – using robust standard errors and individual post-stratification weights provided to account for unequal selection probabilities and differential non-response – the lack of variables delineating PSUs and strata in the Harmonised LASI dataset mean that the effects of clustering were not fully accounted for. Not accounting for PSUs is likely to have underestimated standard errors, whereas not accounting for stratification may have resulted in overestimated standard errors.

Overall, our findings align with growing evidence from India and other low- and middle-income countries showing an association between unclean cooking fuel use and increased depression symptoms [25–28, 31, 48–50]. Using a larger sample of the LASI data that includes middle-aged adults, not only those aged 60 and above, our study extends upon previous work [32].

In contrast to studies from China reporting stronger effects among women and older age groups [51], we found no evidence of effect modification by categorical age or sex, possibly reflecting differences in exposure patterns. Interestingly, we observed a stronger association among individuals belonging to no/other castes (most privileged), followed by Scheduled Castes (most disadvantaged), although these findings should be interpreted cautiously.

Several mechanisms could explain the stronger association observed in urban areas. The heightened effect of unclean cooking fuel use on depression symptoms may reflect higher levels of outdoor air pollution, overcrowding, and poor ventilation. Indeed, there may be residual confounding by these unmeasured variables that we could not account for; previous studies have found kitchen ventilation to be a likely confounder and effect modifier of the association between solid fuel use and depression symptoms [52, 53]. This finding may also reflect the adverse health effects of rapid, unplanned urbanisation in India, concurrent with expansion of urban slums, wherein exploitative or casual labour arrangements disproportionately affect those belonging to marginalised castes, perpetuating the cycle of immiseration, insecure tenure, and poor health [54]. However, the relatively low prevalence of unclean fuel use in urban areas (12.8%) and the small number of exposed individuals with depression symptoms (n = 931) limit the public health implications of this subgroup finding.

Future research should prioritise direct measurement of indoor air pollution, through personal exposure monitoring and household-level PM_2.5_ measurements, to more precisely understand how varying durations and concentrations of pollutant exposure influence depression symptoms. Seasonal variation should also be considered; for example, during winter months, low temperatures limit the dispersal of pollutants, increasing outdoor air pollution in urban areas, which diffuses into households; in turn, individual-level exposure to indoor air pollution may be greater due to more time spent inside [55, 56]. The Averted Mortality and Disability Adjusted Life Years (ADALYs) methodology is the gold standard for quantifying the health benefits of clean cooking practices through measurement of continuous exposure to household-level PM_2.5_ concentrations, before and after implementing an intervention such as cleaner fuels or improved cookstoves [57].

Our study was restricted to data collected at only one time point. Longitudinal data, including subsequent waves of LASI, will be important to establish any temporal association and potentially draw more meaningful conclusions about causality between exposure to unclean cooking fuels and depression symptoms in India, where current evidence is sparse, and allow cross-national comparisons

Exposure to polluting compounds such as black carbon and PM_2.5_ has been associated with elevated circulating biomarkers of oxidative stress and inflammation, including C-reactive protein and cortisol [23, 58], which have been implicated in depression aetiology [19]. Our ability to investigate potential mediating pathways, such as systemic inflammation, was limited by the lack of relevant biomarker data. Future studies integrating biomarker measurements would help to elucidate the biological mechanisms and mediatory pathways linking household air pollution exposure to mental health outcomes.

In conclusion, our study provides evidence that use of unclean cooking fuels is associated with higher odds of depression symptoms among adults aged 45 years and older in India. While the observed effect size is modest, these findings suggest that indoor air pollution may be a potentially important, modifiable environmental determinant of poor mental health in low- and middle-income settings. Improvements in clean cooking fuel access may therefore confer mental health benefits alongside established physical health benefits.

## Financial Support

This research received no specific grant from any funding agency, commercial or not-for-profit sectors. CWG is supported by a Wellcome Career Development Award (225868/Z/22/Z).

## Competing Interests

None.

## Ethical Standards

Ethical approval for this secondary data analysis was granted by the London School of Hygiene and Tropical Medicine MSc Research Ethics Committee (Ref 30880). Ethical approval for conducting the original LASI study and the use of human participants was granted by the following collaborating organisations: Indian Council of Medical Research (F.No.T.21012/07/2012-NCD), Delhi; International Institute for Population Sciences Institutional Review Board (IRB) (Sr. No. 12/1054), Mumbai; Harvard T.H. Chan School of Public Health IRB (CR-16715-10), Boston; University of Southern California IRB (UP-CG-14_00005), Los Angeles; Indian Council of Medical Research (F.No.T.21012/07/2012-NCD), Pune; and Regional Geriatric Centres IRB, Ministry of Health and Family Welfare. LASI enumerators obtained informed consent from respondents prior to data collection. The Harmonised LASI dataset used in this study is publicly available in anonymised form and was accessed for research purposes between July and September 2024. The authors did not have access to information that could identify individual participants at any stage of the analysis. All methods were conducted in accordance with relevant guidelines and regulations.

## Availability of Data and Materials

LASI Wave 1 2017–18 data and related documentation, including fact sheets, national report and executive summary, are publicly available and can be accessed from the website of the Gateway to Global Aging Data (g2aging.org).

## Author contribution statement

CWG conceived the study. CWG, KEM, GRGL, and KM designed the study. KM conducted the statistical analysis. KM, GRGL, and SDR wrote the first draft of the manuscript. All authors contributed to the data interpretation and critical revision for important intellectual content. All authors approved the final version submitted.

## Data Availability

http://g2aging.org

## Supporting information

**S1 Table. CES-D-10 depression questionnaire**.

**S1 Fig. Directed acyclic graph (DAG) conceptualising the association between the exposure (use of unclean cooking fuel), outcome (depression symptoms), measured and unmeasured confounders, and mediators on the causal pathways.**

**S2 Table. Covariate selection.**

**S2 Fig. Flow chart for inclusion and exclusion into sample population for analyses on unclean cooking fuel usage and depression symptoms.**

**S3 Table. Unweighted characteristics of LASI respondents included (N=62,650) and excluded (N=9,612) in the final study population by cooking fuel status.**

## Notes

### Competing Interest Statement

The authors have declared no competing interest.

### Funding Statement

This research did not receive any specific grant from any funding agency, commercial or not-for-profit sectors. CWG is supported by a Wellcome Career Development Award (225868/Z/22/Z).

