## Supplementary Table 1 for "The association between household use of unclean cooking fuels and depression symptoms among older adults in India: a cross-sectional study"

### **Supporting Table 1 : CES-D-10 depression questionnaire**

|  | **During the past week** | | | |
| --- | --- | --- | --- | --- |
|  | **Rarely or never (less than 1 day)** | **Sometimes**  **(1-2 days)** | **Often**  **(3-4 days)** | **Most or all of the time**  **(5-7 days)** |
| **Had trouble concentrating** | 0 | 1 | 2 | 3 |
| **Felt depressed** | 0 | 1 | 2 | 3 |
| **Everything an effort** | 0 | 1 | 2 | 3 |
| **Felt tired or low energy** | 0 | 1 | 2 | 3 |
| **Was happy*** | 3 | 2 | 1 | 0 |
| **Felt lonely** | 0 | 1 | 2 | 3 |
| **Felt overall satisfied*** | 3 | 2 | 1 | 0 |
| **Felt hopeful about the future*** | 3 | 2 | 1 | 0 |
| **Bothered by little things** | 0 | 1 | 2 | 3 |
| **Felt afraid of something** | 0 | 1 | 2 | 3 |
| *Coding reversed for the three questions ascertaining positive feelings | | | | |
