## Supplementary Table 2 for "The association between household use of unclean cooking fuels and depression symptoms among older adults in India: a cross-sectional study"

**Supporting Table 2: Covariate selection**

| **Construct** | **Covariate** | **Covariate** | **Rationale** | **Description** |
| --- | --- | --- | --- | --- |
|  | **Age** | **Confounder**  **Effect modifier** | Prevalence of depression is highest among older adults (those aged 60 and above) [1]. | Since age was measured as a continuous variable in the LASI survey, we recoded it into three categories of 45 to 59 years, 60 to 74 years, and 75 years and above, that we determined to be biologically meaningful categorisations of middle age, old age, and oldest age respectively based upon previous literature [1], to generate strata with adequate statistical power to investigate effect modification in secondary analyses. |
|  | **Sex** | **Confounder**  **Effect modifier** | Women are disproportionately exposed to indoor air pollution as they spend more time at home than men [2, 3]. Furthermore, the prevalence of depression is higher in women compared to men in India [1]. Widowed, elderly women, and those with less education and low income have highest risk of depression [4-6]. | Male or female |
|  | **Marital status** | **Confounder** | Loneliness in older age and widowhood increases susceptibility to mental illnesses such as depression, as well as cognitive impairment and neurodegenerative disease [7]. | We recoded marital status into two categories of married/partnered and separated/divorced/widowed. |
|  | **Rural/urban** | **Confounder**  **Effect modifier** | Large rural-urban disparities in access to non-polluting fuels mean that rural households are more likely to use unclean cooking fuels [8]. Although there is an established association between urbanicity and mental disorders in HICs, evidence for this remains equivocal in India, wherein the urban slum concretises the social and spatial sequestering of classes [9-11]. | Rural or urban |
| **Socioeconomic vulnerability** | **Caste** | **Confounder**  **Effect modifier** | Those belonging to socially disadvantaged castes (SC/ST/OBC) are more likely to be exposed to pollutants and toxins, owing to spatial segregation. Furthermore, caste disadvantage is correlated with poverty and poor housing conditions such as lack of improved sanitation or owning a water facility inside dwelling [12]. Caste discrimination is also associated with poor mental health outcomes. | Scheduled Castes, Scheduled Tribes, Other Backward Castes, or no/other caste |
|  | **Literacy** | **Confounder** | Literacy and education are strong predictors of adoption of alternative, clean cooking fuels such as LPG [1, 6], and those with less education are more likely to use polluting fuels due to lack of awareness about harms [13]. | Illiterate or literate |
|  | **Education** | **Confounder** | Those with less education are more likely to use polluting fuels and have higher risk of depression [4, 14]. | We recoded education level into five categories of none/below primary school, primary school, middle school, secondary school, and graduation/post-graduation/diploma/certificate/professional course. |
|  | **Employment** | **Confounder** | Lack of employment and income is often associated with poor mental health and depressive symptoms. | Unemployed or employed |
|  | **Above/below poverty line** | **Confounder** | Use of unclean cooking fuels is often correlated with poverty, low income, poor housing conditions, and illiteracy. Socioeconomic disadvantage and poverty are associated with increased risk of depression [15, 16]. | Above or below the International Poverty Line (household income $1.90 per person per day) |
| **Housing conditions** | **Improved sanitation** | **Confounder** | Lack of access to clean water and improved sanitation have been found to be associated with increased odds of depression in India [17] and other LMICs [18, 19]. | Improved sanitation refers to flush or pour flush toilet connected to a piped sewer system, septic tank, pit latrine, or somewhere else. Unimproved sanitation refers to a flush or pour flush toilet that flushes to somewhere besides a piped sewer system, septic tank, or pit latrine; use of other or no facility; use of open space or field; or if the household shares their toilet facility with any other households, regardless of the type of toilet facility. |
|  | **Electricity** | **Confounder** | Adoption of cleaner cooking fuels is associated with income and expenditure; households with access to electricity are more willing to pay to adopt cleaner cooking fuels [20]. | Housing respondent reports whether or not the residence has electricity. |
|  | **Housing materials** | **Confounder** | Housing type and materials (permanent/temporary) have been found to be associated with adverse health effects of indoor air pollution in women and children [21]. Furthermore, a recent analysis of the NFHS-V investigating the factors associated with unclean cooking fuel usage found that, overwhelmingly, rural households, Scheduled Tribes households, and those with kutcha (temporary) housing materials were most likely to use polluting fuels [22]. | Indicates whether the type of housing material is pucca (permanent) or kutcha (temporary/semi-permanent). Pucca means that the roof, wall and floor are all made of permanent materials, defined to include cement, concrete, oven-burnt bricks, hollow cement or ash bricks, stone, metal sheets, timber, tiles, slate, asbestos cement sheet, veneer, plywood, artificial wood. Kutcha is defined to include grass, thatch, palm leaf, bamboo, plastic, polythene sheeting, mud, dung, palm, un-burnt brick, wood, or handmade tiles. |
|  | **Separate bedrooms** | **Confounder** | Housing environment indicators such as noise, poor ventilation, and crowding are detrimental to mental health. For example, a longitudinal study investigating the effects of long-term exposure to polluting fuels in older adults in China found that those living in small houses or houses with small number of rooms had increased risk of depression if they used unclean fuels for heating or cooking [23]. | Coded as 1 if the housing respondent reports more than one room in their home (excluding bathrooms, balconies, passages, and kitchens) and one or more of their rooms is a bedroom. Coded as 0 if the housing respondent reports only one room in their home or more than one room but zero bedrooms. |
|  | **Water facility inside own dwelling** | **Confounder** | Lack of access to clean water is correlated with socioeconomic deprivation and poor mental health outcomes, including increased risk of depression, in India and other settings [24]. | Coded as 1 if the housing respondent reports that their main source of drinking water is in their own dwelling or own yard/plot and their main source of water is piped water, public tap/standpipe, tube well or bore well, dug well, spring water, tanker, cart with small tank, surface water, or another source. Coded as 0 if the housing respondent reports that their main source of drinking water is outside dwelling or bottled water/pouch water or rainwater. |
