## Supplementary Table 3 for "The association between household use of unclean cooking fuels and depression symptoms among older adults in India: a cross-sectional study"

**Supporting Table 3: Unweighted characteristics of LASI respondents included (N=62,650) and excluded (N=9,612) in the final study population by cooking fuel status**

|  | **Uses clean cooking fuel (N=38,376)** | | **Uses unclean cooking fuel (N=32,504)** | |
| --- | --- | --- | --- | --- |
|  | **Excluded** | **Included (N=33,653)** | **Excluded** | **Included (N=28,997)** |
|  | **(N=4,723)** |  | **(N=3,507)** |  |
| **Individual characteristics** | | | | |
| **Age (years) (median [IQR])** | 42.00 [39.00-44.00] | 58.00 [50.00-66.00] | 42.00 [39.00-44.00] | 59.00 [51.00-67.00] |
| **Gender** |  |  |  |  |
| Male | 517 (11.0) | 15,639 (46.5) | 421 (12.00) | 13,423 (46.3) |
| Female | 4,206 (89.0) | 18,014 (53.5) | 3,086 (88.00) | 15,574 (53.7) |
| **Caste** |  |  |  |  |
| Scheduled caste | 703 (14.9) | 4,929 (14.7) | 634 (18.1) | 5,565 (19.2) |
| Scheduled tribe | 499 (10.6) | 3,332 (9.9) | 920 (26.2) | 7,595 (26.2) |
| Other backward caste | 1,886 (39.9) | 13,296 (39.5) | 1,226 (35.0) | 10,287 (35.5) |
| No caste/other | 1,576 (33.4) | 11,777 (35.0) | 697 (19.9) | 5,397 (18.6) |
| *Missing* | 59 (1.2) | 319 (0.9) | 30 (0.9) | 153 (0.5) |
| **Literacy** |  |  |  |  |
| Literate | 3,394 (71.9) | 21,730 (64.6) | 1,518 (43.3) | 10,654 (36.7) |
| Illiterate | 1,326 (28.1) | 11,920 (35.4) | 1,987 (56.7) | 18,339 (63.2) |
| *Missing* | 3 (0.1) | 3 (0.0) | 2 (0.1) | 4 (0.01) |
| **Education** |  |  |  |  |
| None/below primary | 1,650 (34.9) | 15,117 (44.9) | 2,314 (66.0) | 21,450 (74.0) |
| Primary | 765 (16.2) | 4,926 (14.6) | 464 (13.2) | 3,315 (11.4) |
| Middle school | 703 (14.9) | 3,904 (11.6) | 394 (11.2) | 2,107 (7.3) |
| Secondary | 1,149 (24.3) | 6,473 (19.2) | 309 (8.8) | 1,807 (6.2) |
| Diploma/grad/post-grad | 455 (9.6) | 3,232 (9.6) | 25 (0.7) | 318 (1.1) |
| *Missing* | 0 (0.0) | 1 (0.0) | 1 (0.0) | 0 (0.0) |
| **Employment** |  |  |  |  |
| Yes | 1,710 (36.2) | 15,123 (44.9) | 1,735 (49.5) | 16,004 (55.2) |
| No | 3,003 (63.6) | 18,529 (55.1) | 1,766 (50.4) | 12,993 (44.8) |
| *Missing* | 10 (0.2) | 1 (0.0) | 6 (0.2) | 0 (0.0) |
| **Marital status** |  |  |  |  |
| Married/partnered | 4,414 (93.5) | 25,544 (75.9) | 3,194 (91.1) | 21,725 (74.9) |
| Separated/divorced/widow | 307 (6.5) | 8,107 (24.1) | 312 (8.9) | 7,272 (25.1) |
| *Missing* | 2 (0.04) | 2 (0.01) | 1 (0.0) | 0 (0.0) |
| **Household characteristics** | | | | |
| **International poverty line** |  |  |  |  |
| Above | 4,399 (93.1) | 30,912 (91.9) | 2,531 (72.2) | 21,102 (72.8) |
| Below | 324 (6.9) | 2,741 (8.1) | 976 (27.8) | 7,893 (27.2) |
| *Missing* | 0 (0.0) | 0 (0.0) | 0 (0.0) | 2 (0.0) |
| **Rural/urban residence** |  |  |  |  |
| Rural | 1,956 (41.4) | 14,930 (44.4) | 3,121 (89.0) | 25,848 (89.1) |
| Urban | 2,767 (58.6) | 18,723 (55.6) | 386 (11.0) | 3,149 (10.9) |
| **Improved sanitation** |  |  |  |  |
| Yes | 3,840 (81.3) | 28,016 (83.2) | 2,199 (62.7) | 17,213 (59.4) |
| No | 876 (18.6) | 5,619 (16.7) | 1,307 (37.3) | 11,781 (40.6) |
| *Missing* | 7 (0.1) | 18 (0.1) | 1 (0.0) | 3 (0.0) |
| **Water facility inside dwelling** |  |  |  |  |
| Yes | 3,618 (76.6) | 26,473 (78.7) | 2,106 (60.0) | 17,162 (59.2) |
| No | 1,105 (23.4) | 7,174 (21.3) | 1,398 (39.9) | 11,816 (40.7) |
| *Missing* | 0 (0.0) | 6 (0.0) | 3 (0.1) | 19 (0.1) |
| **Housing materials** |  |  |  |  |
| Permanent (pucca) | 3,298 (69.8) | 24,360 (72.4) | 1,139 (32.5) | 9,554 (33.0) |
| Temporary (kutcha) | 1,399 (29.6) | 9,231 (27.4) | 2,358 (67.2) | 19,406 (66.9) |
| *Missing* | 26 (0.6) | 62 (0.2) | 10 (0.3) | 37 (0.1) |
| **Separate bedrooms** |  |  |  |  |
| Yes | 3,970 (84.1) | 29,253 (86.9) | 2,796 (79.7) | 22,933 (79.1) |
| No | 752 (15.9) | 4,397 (13.1) | 711 (20.23) | 6,062 (20.9) |
| *Missing* | 1 (0.0) | 3 (0.0) | 0 (0.0) | 2 (0.01) |
| **Electricity** |  |  |  |  |
| Yes | 4,676 (99.0) | 33,287 (98.9) | 3,131 (89.3) | 25,620 (88.3) |
| No | 47 (1.0) | 366 (1.1) | 376 (10.7) | 3,377 (11.7) |
