## Supplementary figures and images for "The association between household use of unclean cooking fuels and depression symptoms among older adults in India: a cross-sectional study"

### Supplementary Figure 1

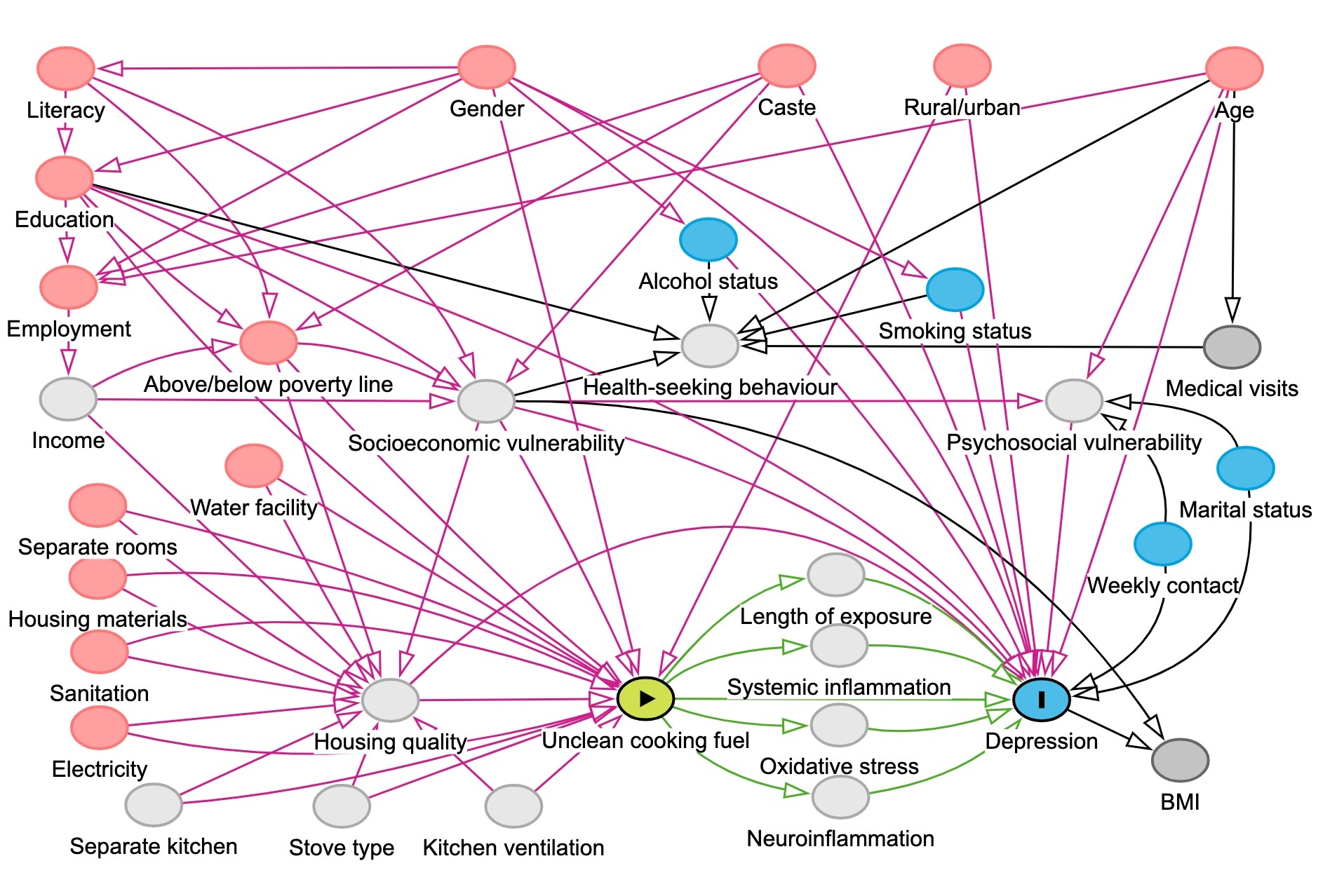

### Supplementary Figure 2

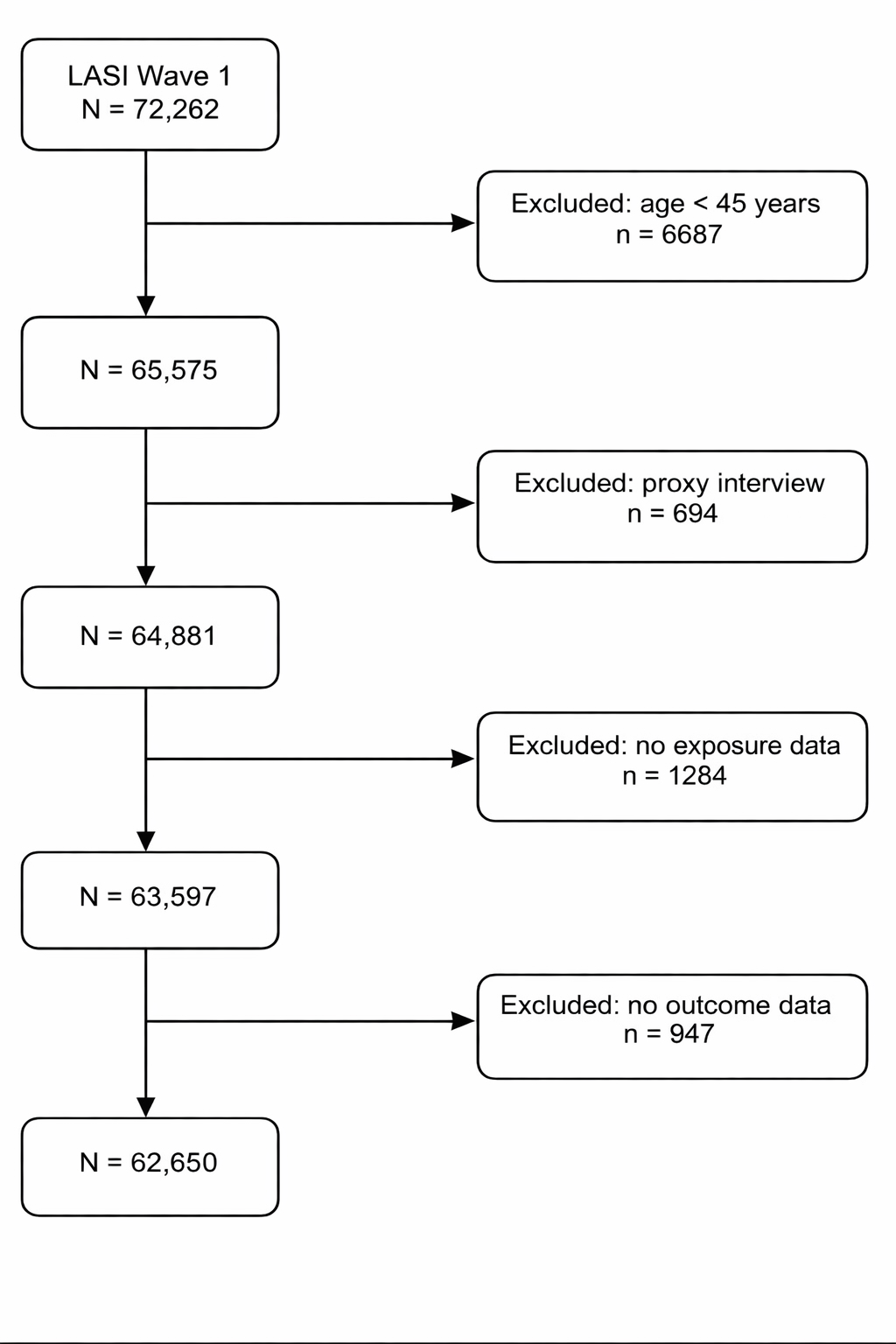
